# Levels of produced antibodies after vaccination with mRNA vaccine; effect of previous infection with SARS-CoV-2

**DOI:** 10.1101/2021.04.05.21254934

**Authors:** Theocharis Konstantinidis, Stavroula Zisaki, Ioannis Mitroulis, Eleni Konstantinidou, Eftychia G. Kontekaki, Gioulia Romanidoui, Alexandros Karvelas, Ioanna Nanousi, Leonidas Lazidis, Dimitrios Cassimos, Christina Tsigalou, George Martinis, Maria Panopoulou

## Abstract

The aim of this study was to estimate the immunogenic effect of mRNA vaccine against SARS-CoV-2. This study included 510 participants who received mRNA vaccine. The measurement of anti-Covid-19 antibodies was performed using the Abbott SARS-CoV-2 IgG quantitative assay (Abbott). Overall, mean title of anti-Spike antibodies was 19319.2±1787.5 AU/ml. Vaccination induced a robust immunogenic response in previous infected with SARS-CoV-2 compared. Additionally, individuals that were asymptomatic after vaccination produced lower levels of antibodies compared to feverish individuals. In conclusion, remarkable high level of anti-Spike Covid-19 antibodies was found after vaccination.

## 1. Introduction

The novel Coronavirus Disease (COVID-19) due to the Severe Acute Respiratory Syndrome Coronavirus 2 (SARS CoV-2) is an emerging global health problem that reported for the first time in Wuhan, China in December 2019 [1]. Due to person-to-person transmission, the infection was rapidly spread worldwide.

Vaccination is a safe, and effective method of protecting against infectious diseases. Vaccines train the immune response to recognize and neutralize an infectious agent. On account of the long-standing vaccination programs during childhood and adults large epidemics of infectious diseases have reduced worldwide [2].

The insufficient the SARS-CoV-2 pandemic control require rapid development of vaccines. There are lots of anti-SARS-CoV-2 vaccine candidates, based on several different mechanisms of action are currently in development or finished phase III trial [3,4]. Since December 2020, several vaccines have been authorized worldwide. In Greece, vaccination was started from Healthcare professionals, and on 13 March 2021, 1.283.472 doses of vaccines were used [5].

The aim of this study was to estimate the seroprevalence of SARS-CoV-2 antibodies among persons who take 2 doses of mRNA vaccine in Thrace region and explore risk factors for any adverse events.

## 2. Materials and Methods

### 2.1. Study design

This study was performed at the University General Hospital of Alexandroupolis and Democritus University of Thrace in Alexandroupolis, Greece. The study protocol was approved by the local committee of ethics and deontology in accordance with the Declaration of Helsinki (Number 1070/11-01-2021).

### 2.2. Study population

This study included 510 persons, both uninfected (n=487) and previously infected persons (n=23) with confirmed COVID-19 by RT-PCR, occurring 2 to 3.5 months prior to vaccination. All persons were vaccinated with mRNA (BNT162b2 Pfizer/BioNTech) vaccine. Serum samples were collected before vaccination and 1 month after second dose. The period of sampling was from 10 February to 16 March 2021.

### 2.3. IgG Testing

The measurement of anti-Covid-19 antibodies was performed using the Abbott Architect i1000SR instrument (Abbott Diagnostics, IL, USA) using the Abbott SARS-CoV-2 IgG quantitative kit and following manufacturer’s instructions. The assay is a chemiluminescent microparticle immunoassay for qualitative detection of anti-SARS-CoV-2 Abs type IgG against the CoV-2 Spike protein (Sp) in human serum. Quantitative results >50 AU/ml are reported as positive in accordance with the Abbott-determined positivity cutoff of 50 AU/ml.

### 2.4. Statistical Analysis

Continuous variables were presented as mean ± standard deviation (SD) for normally distributed data. The counting data were expressed by rate (%). Mann Whitney U test was used for independent samples and Wilcoxon test for paired sample analysis. The P value < 0.05 indicated a statistically significant difference.

## 3. Results

### 3.1. Safety assessment

Overall, 510 persons were enrolled in the study. The demographic data is presented in Table 1. Summary data (numbers and percentages) for participants with any adverse events shown that the most common were local adverse events; the injection-site event was pain after injection were noted in 114 participants (22.5%) after the first dose and/or the second dose, and they resolved over the following 1 to 5 days. The most common system adverse events were fever-95 (18.6%), headache-78 (15.3%), myalgias-68 (13.3%), arthralgia-12 (2%), fatigue-57 (11.2%), and lymphadenopathy-22 (4.3%) (Table 1). The severe adverse events leading to discontinuation of the second dose injections were recorded in 3/510 (0.59%) participants, who present severe allergic reactions. Only one woman developed delayed hypersensitivity reaction. Initially started as macular on 4^th^ day after vaccination, but subsequently developed maculopapular lesions with symmetrical distribution on the extremities.

**Table 1.** Demographical, clinical, and adverse events data of study population CVD-Cardiovascular diseases

|  | **Non-Infected**<br>**n= 487** | **COVID-19**<br>**n= 23** | **Overall**<br>**n= 510** |
| --- | --- | --- | --- |
| **Age** | 48.4±2.5 | 47.1±2.3 | 47.5±2.5 |
| **Male** | 147 (30.2%) | 6 (26.1%) | 153 (30%) |
| **Female** | 340 (69.8%) | 17 (73.9%) | 357 (70%) |
| **Occupational Risk** |  |  |  |
| **Healthcare Workers** | 457 (93,8%) | 23 (100%) | 480 (94%) |
| **Underlying disease** |  |  |  |
| **CVD** | 18 (3,7%) | 1 (4.3%) | 19 (3,7 |
| **Hypertension** | 22 (4,5%) | 2 (8.3%) | 24 (4,7%) |
| **Diabetes mellitus** | 17 (3,5%) | 2 (8.3%) | 19 (3,7%) |
| **Cancer** | 6 (1,2%) | - | 6 (1,2%) |
| **Autoimmune diseases** | 15 (3.1%) | - | 15 (2.9%) |
| **Adverse events** |  |  |  |
| **No Adverse Events** | 184 (38,1%) | 5 (21%) | 189 (37%) |
| **Solicited Local** |  |  |  |
| **Pain** | 106 (21,9%) | 8 (34,8%) | 114 (22,5%) |
| **Swelling** |  |  |  |
| **Lymphadenopathy** | 20 (4,1%) | 2 (8,7%) | 22 (4,3%) |
| **Supraclavicular** | 3 (0,6%) | - | 3 (0,59%) |
| **Axillary** | 17 (3,5%) | 2 (8,7%) | 19 (3,75%) |
| **Systemic Adverse Events** |  |  |  |
| **Fever** | 90 (18.5%) | 5 (21.7%) | 95 (18,2%) |
| **Shiver** | 79 (16.2%) | 5 (21.7%) | 84 (16.5%) |
| **Headache** | 74 (15.9%) | 4 (17.4%) | 78 (15.3%) |
| **Fatigue** | 54 (11.1%) | 3 (13%) | 57 (11.2%) |
| **Nausea/vomiting** | 15 (3.1%) | - | 15 (2.9%) |
| **Myalgia** | 64 (13.1%) | 4 (17.4%) | 68 (13.3%) |
| **Arthralgia** | 8 (1.6%) | 4 (17.4%) | 12 (2.4%) |
| **Hypersensitivity** |  |  |  |
| **Type IV hypersensitivity reaction** | 1 (0,2%) | - | 1 (0,19%) |
| **Edema** | 3 (0,6%) | - | 3 (0,59%) |

### 3.2. The immunogenic effect

Overall, the immunization with administration of two doses were completed in 507 participants (99,4%). Anti-Spike SARS-CoV-2-IgG antibodies were detected in 508 out of 510 (99,6 %) with mean value 19319.2±1787.5 AU/ml. Participants who suffered from COVID-19 had higher levels of anti-Spike antibodies in comparison to non-infected 25599.5±10646.8 vs 19221.3±1803.66, p=0.049, respectively (Figure 1A). Moreover, COVID-19 patients had developed higher levels of anti-Spike antibodies after vaccination 593.7±379.2 vs 25599.5±10646.8 AU/ml, p<0.00001 (Figure 1B). Patients with fever developed higher titer of anti-Spike Abs in comparison to asymptomatic participants (28899.6±4831.01 vs 14685.9±214.1 p< 0,00001) Figure 1C. Moreover, patients with autoimmune disorders had lower titer of anti-Spike Abs than general population in a statistically significant manner 6311.18±557.1 vs 19319.2±1787.5 AU/ml.

**Figure 1.**
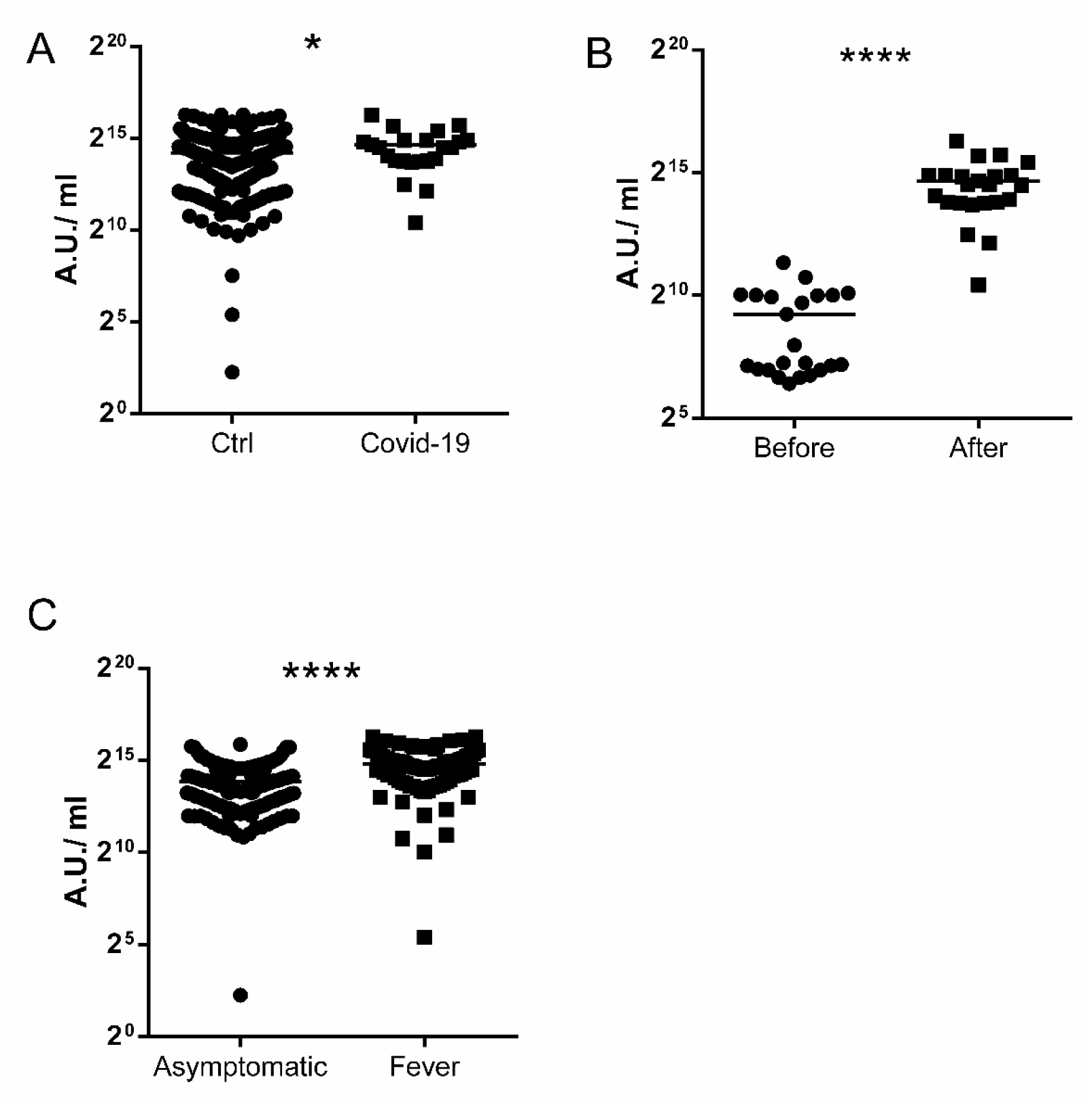
The immunogenic effect of mRNA vaccine. A- Levels of antibodies after vaccination. Comparison of controls vs COVID-19 patients B- Levels of antibodies in COVID-19 patients before and after vaccination C- Levels of antibodies after vaccination in asymptomatic persons vs systemic adverse events (fever) persons The date is presenting as log2. *-p=0.049, ****-p<0.0001

## 4. Discussion

This study provides evidence of short-term efficacy of the mRNA vaccine in preventing symptomatic SARS-CoV-2 infection in an adult population in Thrace region, Greece. Overall, in this study, only three severe adverse events after receipt of the first vaccine that led to postpone the second dose has been reported. The adverse events after receipt of Pfizer/BioNTech COVID-19 vaccine in the United States, were reported in 4,393 (0.2%) cases. Among these, cases of severe allergic reaction, including anaphylaxis. were recorded [6]. The cases of acute onset of a single lymphadenopathy (supraclavicular or Axillary) after intramuscular administration of an mRNA-based COVID-19 vaccine, recorded in 20 participants in this study. These results are in line with previously reported by O R Mitchell et al., and Fernández-Prada et al [7,8].

The mean value of anti-SARS-CoV-2 Spike protein in patients with autoimmune disorders in this study was lower than in general population (6311.18±557.1 vs 19319.2±1787.5 AU/ml). The data on specific COVID-19 vaccine response in patients under immunosuppressive therapy is poorly until now. Immunosuppressive therapy in patients with autoimmune disorders or transplantation may impair vaccine responses. This data were previously shown on vaccination of immunosuppressive patients predominantly focus on influenza and pneumococcal vaccines [9]. A limitations of the study is the relatively small group of vaccinated patients after SARS CoV infection.

## Data Availability

The data that support the findings of this study are available from the corresponding author, upon reasonable request.

## Author Contributions

Conceptualization, methodology, and validation, T..K..; laboratory investigation, S.Z., E.K., E.G.K. and I.N.; clinical data curation, I.M., G.R., and D.C.; writing— original draft preparation, T.K. I.M; writing—review and editing, C.T., D.C, T.K. and I.M; supervision, G.M., and M.P.

All authors have read and agreed to the published version of the manuscript.

